# The UK COVID-19 furlough scheme and associations with smoking, alcohol consumption and vaping: evidence from 8 UK longitudinal population surveys

**DOI:** 10.1101/2021.10.28.21265593

**Authors:** Michael J. Green, Jane Maddock, Giorgio Di Gessa, Bożena Wielgoszewska, Sam Parsons, Gareth J Griffith, Jazz Croft, Anna J. Stevenson, Charlotte F. Huggins, Charlotte Booth, Jacques Wels, Richard J. Silverwood, Praveetha Patalay, Alun D. Hughes, Nishi Chaturvedi, Laura D Howe, Emla Fitzsimons, Srinivasa Vittal Katikireddi, George B. Ploubidis

## Abstract

**Background:** Disruptions to employment status can impact smoking and alcohol consumption. During the COVID-19 pandemic, the UK implemented a furlough scheme to prevent job loss. We examine how furlough was associated with participants’ smoking, vaping and alcohol consumption behaviours in the early stages of the pandemic.

**Methods:** Data were from 27,841 participants in eight UK adult longitudinal surveys. Participants self-reported employment status and current smoking, current vaping and drinking alcohol (>4 days/week or 5+ drinks per typical occasion) both before and during the pandemic (April-July 2020). Risk ratios were estimated within each study using modified Poisson regression, adjusting for a range of potential confounders, including pre-pandemic behaviour. Findings were synthesised using random effects meta-analysis. Sub-group analyses were used to identify whether associations differed by gender, age or education.

**Results:** Compared to stable employment, neither furlough, no longer being employed, nor stable unemployment were associated with smoking, vaping or drinking, following adjustment for pre-pandemic characteristics. However, some sex differences in these associations were observed, with stable unemployment associated with smoking for women (ARR=1.35; 95% CI: 1.00-1.82; I^2^: 47%) but not men (0.84; 95% CI: 0.67-1.05; I^2^: 0%). No longer being employed was associated with vaping among women (ARR=2.74; 95% CI: 1.59-4.72; I^2^: 0%) but not men (ARR=1.25; 95% CI: 0.83-1.87; I^2^: 0%). There was little indication of associations with drinking differing by age, gender or education.

**Conclusions:** We found no clear evidence of furlough or unemployment having adverse impacts on smoking, vaping or drinking behaviours during the early stages of the COVID-19 pandemic in the UK, with differences in risk compared to those who remained employed largely explained by pre-pandemic characteristics.

## Introduction

The COVID-19 pandemic and the measures many countries have implemented to contain the spread of the virus represent a major societal disruption (1, 2). For example, UK restrictions came into effect from 23^rd^ March 2020 requiring staying at home (except for limited purposes such as shopping or essential work), and closure of certain businesses (e.g. retail and leisure) (3). COVID-19 has been especially disruptive to economic activity, with many stopping work, losing income or reducing hours (4–6). Many high-income countries have introduced measures to maintain employment during the pandemic. In particular, furlough schemes have replaced lost income for workers temporarily unable to carry out their job. The UK introduced a furlough policy, the Coronavirus Job Retention Scheme (CJRS), providing 80% of pay (capped at £2,500 a month) for employees who were unable to work during the pandemic (3). Understanding the impact of this policy is crucial not only to inform the COVID-19 response internationally but also to inform employment policies post-pandemic.

Smoking cigarettes and drinking alcohol seriously damage health and contribute substantially to health inequalities (7–9). COVID-19-related disruptions have the potential for considerable impact on smoking and drinking behaviours, though evidence so far is mixed and suggests heterogeneous impacts (10–14). Employment-related disruptions may be important, as increases in smoking and drinking are often associated with loss of work (15–17). It is unclear whether similar effects would be seen for stopping work as part of a furlough scheme, or during a pandemic, though some evidence already indicates that furlough is associated with increased alcohol intake (14, 18).

Health impacts of the CJRS could differ, for example, by age, gender or socioeconomic position, so we need to understand who is most affected and to what extent this could modify existing inequalities (2, 6, 19). Younger workers, women and low earners tend to work in industries/sectors where people are more likely to lose jobs and/or be furloughed (20). Socioeconomic inequalities in smoking are well known, while evidence is more mixed for drinking (21–24). Both behaviours tend to peak in young adulthood with either cessation or decreases thereafter (25, 26). Additionally, there is evidence that younger generations drink and smoke less (27, 28) and that gender differences (namely that men tend to smoke and drink more) have narrowed (29, 30). Vaping (i.e. use of electronic or e-cigarettes) has also become more common in recent years as a potentially less harmful alternative to smoking (31, 32).

The UK National Core Studies Longitudinal Health and Wellbeing initiative draws together data from multiple UK population-based longitudinal studies using coordinated analysis to answer priority pandemic-related questions. By conducting harmonised primary analyses within each study and pooling results in a meta-analysis, we can provide robust evidence to understand how the pandemic has impacted population health and behaviours, and support efforts to mitigate its effects going forward. Here, we aim to examine how changes in employment status during the pandemic, especially participation in the furlough scheme, are associated with smoking, vaping and drinking behaviours. We further assess whether associations vary by gender, age or education.

## Methods

### Design

#### Participants

Data were drawn from eight UK population studies that conducted surveys both before and during the COVID-19 pandemic. Details of the design, sample frames, current age range, timing of the most recent pre-pandemic and COVID surveys, response rates, and analytical sample size are available in Supplementary File 1.

Five of these were age homogenous birth cohorts (all individuals similar age): the Millennium Cohort Study (MCS; 33, 34); the children of the Avon Longitudinal Study of Parents and Children (ALSPAC-G1; 35); Next Steps (NS, formerly known as the Longitudinal Study of Young People in England; 34, 36); the 1970 British Cohort Study (BCS70; 34, 37); and the 1958 National Child Development Study (NCDS; 34, 38). Three age heterogeneous studies (covering a range of age groups) were also included: Understanding Society (USOC; 39); the English Longitudinal Study of Ageing (ELSA; 40); and Generation Scotland: the Scottish Family Health Study (GS; 41). Finally, the parents of the ALSPAC-G1 cohort were treated as a fourth age heterogeneous study population (ALSPAC-G0).

Analytical samples were restricted to working age (16-66 years, based on the current UK state pension age; 42) and participants who had recorded at least one outcome in a COVID-19 survey between April and July 2020 (the period covered by the first lockdown restrictions in the UK) and had valid covariate data. Most studies were weighted to be representative of their target populations, accounting for sampling design, and differential non-response (34). Weights were not available for GS.

### Measures

Below we describe measurement of all variables. Full details of the questions and coding used within each study are available in Supplementary File 2.

#### Exposure: Employment status change

Employment status change (or stability) was coded in six categories based on employment status both prior to the pandemic and at their first COVID-19 survey (April-July 2020; see Supplementary File 1 for details by study): stable employed (reference); furloughed (i.e. from work to furlough); no longer employed (i.e. from work to non-work); became employed (i.e. from non-work to work); stable unemployed (i.e. unemployed but looking for work at both points); and stable non-employed (i.e. not employed at either point, including education, early retirement, caring for the home, sick or disabled). Given our primary interest in comparing the behaviours of those in stable employment (seen *a priori* as the optimal condition) to those who were furloughed, no longer employed and stable unemployed, we focus on results for these groups (with findings for those who became employed or were in stable non-employment available in Supplementary File 3).

#### Outcomes: Smoking, Vaping and Drinking behaviours

We examined smoking, vaping, and drinking alcohol. Smoking and vaping were based on any current use (yes vs no). Regarding drinking alcohol, measures of frequency (4+ days per week vs less frequently) and quantity per typical drinking occasion (5+ standard alcoholic drinks per occasion vs less) could be coded consistently across most studies, and we created a combined binary variable indicating a high frequency or quantity or both (Supplementary File 6 presents analyses for frequency and quantity measures separately, but results were largely consistent with each other and with the main analyses). Primary analyses were based on studies/outcomes with measures both during and before the pandemic, whereas for some studies (ELSA/ALSPAC/GS) only information on change since the start of the pandemic was available for drinking. Thus, based on reported consumption before and during the pandemic, or on reported changes in behaviour, we created additional dichotomous variables indicating (in comparison to no change or change in the opposite direction): increased smoking (including take-up/relapse/or more cigarettes); decreased smoking (including cessation and fewer cigarettes); increased vaping (including take-up/relapse or more frequent use); decreased vaping (including cessation and less frequent use); increased drinking (increased frequency or more drinks per occasion or both); and decreased drinking (decreased frequency or fewer drinks per occasion or both). All information on behaviours during the pandemic was from surveys conducted during the first UK lockdown between April and July 2020 (inclusive).

#### Confounders and Moderators

Confounders included: sex (female vs male); ethnicity (non-white ethnic minority vs White - including white ethnic minorities); age; education (degree vs no degree); UK nation (i.e. England, Wales, Scotland or Northern Ireland or other); household composition (based on presence of a spouse/partner and presence of children); pre-pandemic psychological distress (indicated by symptoms above thresholds on standard screening scales); pre-pandemic self-assessed health (excellent-good vs fair-poor); and pre-pandemic measures of smoking, vaping and drinking.

We conducted sub-group analyses of the associations by sex, education and age in three categories: 16-29; 30-49; and 50 years or more (age-homogeneous cohorts were included in the relevant age band).

### Analysis

Within each study, each outcome was regressed on employment status change, using a modified Poisson model with robust standard errors that returns risk ratios (RR) for ease of interpretation (43) and to avoid issues related to non-collapsibility of the odds ratio (44). After estimating unadjusted associations, confounder adjustment was undertaken in two steps. First, a “basic” adjustment including sociodemographic characteristics: age (only in age-heterogeneous studies), sex, ethnicity (except the BCS and NCDS cohorts which were nearly entirely white), education, UK nation (except ALSPAC and ELSA which only had participants from a single country), and household composition. Second, a “full” adjustment additionally including pre-pandemic measures of psychological distress, self-rated health and each outcome behaviour. Sub-group analyses by sex, education and age were performed with stratified regressions using the “full” adjustment.

Both stages of adjustment are relevant because our employment change exposure incorporates pre-pandemic employment status, which may have influenced other pre-pandemic characteristics such as mental health, self-rated health, and outcome behaviours (see Supplementary File 4, Figure S1). By not controlling for these pre-pandemic characteristics, the basic adjusted risk ratios may represent both newly acquired behaviour and/or continuation of established (pre-pandemic) behaviour. In contrast, the full adjustment risk ratios block effects via these pre-pandemic characteristics and are interpreted as representing the differential change in health behaviour between exposure groups which is independent of these pre-pandemic characteristics. For the outcomes directly capturing changes in health behaviour, the full adjustment did not include pre-pandemic levels of the behaviour in question as pre-pandemic levels of that behaviour are incorporated within the change outcome. This means that even full adjustment risk ratios estimated for these outcomes may partially reflect associations with pre-pandemic behaviour.

The overall and stratified results from each study were pooled using a random effects meta-analysis with restricted maximum likelihood. Differences in estimates between sub-groups were tested using the subgroup meta-analysis command. Some studies could not contribute estimates for every comparison due to differences in the ages sampled, measures used, and sparsity of data (45). For a small number of exposure levels the number of outcome cases was low (≤2) and therefore estimates were deemed as unreliable and excluded. While such selective exclusion could potentially lead to bias, the low numbers of events mean that the corresponding within-study estimates would be so imprecise that their exclusion is unlikely to lead to considerable bias (see Supplementary File 3 for more details and sensitivity analyses showing that results were robust to different low cell count exclusion thresholds). We report heterogeneity using the I2 statistic: 0% indicates estimates were similar across studies, while values closer to 100% represent greater heterogeneity. All analyses were conducted using Stata 16 or R.

## Results

Analyses included 27,841 individuals from eight studies (see Supplementary File 5, Table S1 for sample characteristics). Figure 1 shows patterns of employment status change from pre-pandemic to those recorded in April-July 2020. Around six in 10 participants in NS, BCS, GS, USOC, and ALSPAC G0 were employed before and during the initial stages of the pandemic, with the younger (MCS) and older studies (ELSA and NCDS) showing lower levels of stable employment. Prevalence of furlough ranged between 8% (GS) and 26% (NS). Across most studies, approximately 3% of participants were no longer employed during the pandemic (7% in ALSPAC G0). Stable unemployment ranged in prevalence between 1% (GS) and 8% (ALSPAC G0). Supplementary File 5 - Table S2 shows how economic activity was patterned by education, sex, and age-groups, with furlough generally more common among younger, female and less educated participants and stable employment especially common among male, more highly educated and middle-aged participants. There were no clear patterns across studies with regard to who was no longer employed during the pandemic, but no longer being employed was consistently rare.

**Figure 1:**
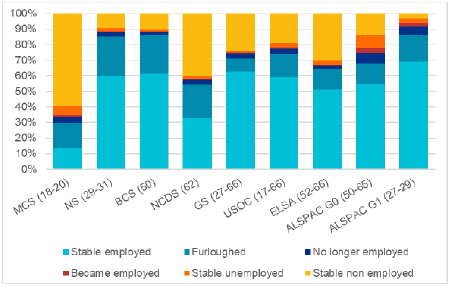
Patterns of employment status change in the initial stages of the pandemic by study. Parentheses show the age range of participants in each study sample at the time of data collection. Weighted data (except GS). Analysis for GS, USOC, and ELSA restricted to participants aged 66 and younger. Supplementary File 2 details the questions asked in each study.

Table 1 shows the prevalence of smoking, vaping and drinking, and of changes from pre-pandemic behaviour, in each of the eight studies. Only USOC and ELSA had comparable data on smoking prevalence both before and during the pandemic, but proportions were similar (in the MCS, NS, BCS and NCDS cohorts, for example, the questions on frequency of pre-pandemic smoking were only asked for those who reported smoking during the pandemic). Vaping was less common than smoking in all studies where it had been recorded and the pre-pandemic prevalence from USOC (7.5%) was similar to that observed during the pandemic (6.6%). Reported changes in smoking and vaping behaviour were relatively rare.

**Table 1.** Percent (and N) distribution of health behaviours and changes during the pandemic by study.

|  | **MCS** | **NS** | **BCS** | **NCDS** | **GS** | **USOC** | **ELSA** | **ALSPAC**<br>**G0** | **ALSPAC**<br>**G1** |
| --- | --- | --- | --- | --- | --- | --- | --- | --- | --- |
| N participants | 2057 | 1579 | 3151 | 4358 | 2604 | 8328 | 2417 | 2072 | 1275 |
| Age/Age range of participants | 18-20 | 29-31 | 50 | 62 | 27-66 | 17-66 | 52-66 | 50-65 | 27-29 |
|  | **% (N)** | **% (N)** | **% (N)** | **% (N)** | **% (N)** | **% (N)** | **% (N)** | **% (N)** | **% (N)** |
| **Smoking and Vaping** |  |  |  |  |  |  |  |  |  |
| Pre-Pandemic Current Smoker | NA | NA | NA | NA | 12.2<br>(317) | 15.5<br>(834) | 14.7<br>(271) | NA | NA |
| Current smoker (during pandemic) | 17.3<br>(292) | 18.0<br>(189) | 16.7<br>(366) | 11.3<br>(356) | 6.2<br>(162) | 14.7<br>(793) | 13.3<br>(240) | NA | NA |
| Smoking more -including relapse and initiation | 5.4<br>(78) | 8.5<br>(76) | 6.5<br>(136) | 2.9<br>(85) | 2.7<br>(71) | 6.3<br>(356) | 4.4<br>(77) | NA | NA |
| Smoking less -including cessation | 6.9<br>(137) | 4.0<br>(46) | 2.5<br>(57) | 1.0<br>(41) | 1.0<br>(24) | 6.8<br>(339) | 4.2<br>(83) | NA | NA |
| Pre-Pandemic Current Vaping | NA | NA | NA | NA | NA | 7.5<br>(490) | NA | NA | NA |
| Current Vaping (during pandemic) | 7.4<br>(122) | 10.2<br>(106) | 9.5<br>(228) | 5.7<br>(189) | 3.3<br>(87) | 6.6<br>(423) | NA | NA | NA |
| Vaping more -including relapse and initiation | 3.0<br>(51) | 5.0<br>(52) | 2.5<br>(67) | 1.0<br>(41) | 1.0<br>(26) | 2.3<br>(124) | NA | NA | NA |
| Vaping less -including cessation | 1.4 | 1.2 | 0.5 | 0.4 | < 1.0 | 3.3 | NA | NA | NA |
|  | (27) | (13) | (15) | (16) | (< 10) | (188) |  |  |  |
| **Alcohol Consumption** |  |  |  |  |  |  |  |  |  |
| Drinks 4+ days a week<br>(pre-pandemic) | 8.6<br>(143) | 6.2<br>(72) | 15<br>(496) | 24<br>(1,145) | NA | 28.2<br>(2314) | NA | NA | NA |
| Drinks 4+ days a week<br>(during the pandemic) | 9.6<br>(151) | 13.8<br>(197) | 27.0<br>(911) | 29.7<br>(297) | NA | 51.7<br>(4272) | NA | NA | NA |
| Drinks more frequently | 17.4<br>(351) | 30.5<br>(492) | 25.3<br>(883) | 15.4<br>(707) | NA | 34.6<br>(3009) | NA | NA | NA |
| Drinks less frequently | 36.2<br>(786) | 14.1<br>(222) | 8.0<br>(242) | 12.4<br>(519) | NA | 24.4<br>(1904) | NA | NA | NA |
| Drinks 5+ drinks per occasion<br>(pre-pandemic) | 30.8<br>(687) | 12<br>(180) | 12<br>(329) | 9.3<br>(372) | NA | 16.8<br>(1040) | NA | NA | NA |
| Drinks 5+ drinks per occasion<br>(during the pandemic) | 10.6<br>(171) | 8.6<br>(102) | 11.7<br>(325) | 9.0<br>(333) | NA | 6.9<br>(485) | NA | NA | NA |
| More drinks per occasion | 17.4<br>(351) | 30.5<br>(492) | 25.3<br>(883) | 15.4<br>(707) | NA | 9.4<br>(743) | NA | NA | NA |
| Fewer drinks per occasion | 46.5<br>(782) | 20.2<br>(239) | 10.4<br>(242) | 9.8<br>(297) | NA | 36.6<br>(2695) | NA | NA | NA |
| Drinks 4+ d/week or 5+ drinks per<br>occasion<br>(pre-pandemic) | 35<br>(748) | 17.9<br>(247) | 24.4<br>(742) | 28.9<br>(1325) | NA | 42.8<br>(3200) | NA | NA | NA |
| Drinks 4+ d/week or 5+ drinks per<br>occasion | 18.2<br>(282) | 21.0<br>(273) | 33.4<br>(1054) | 34.8<br>(1507) | NA | 55.8<br>(4536) | NA | NA | NA |
| (during the pandemic) |  |  |  |  |  |  |  |  |  |
| Drinks more/more frequently | 23.7<br>(396) | 36.7<br>(541) | 31.7<br>(1025) | 21.1<br>(889) | 26.3<br>(685) | 38.4<br>(3329) | 18.1<br>(455) | 38.1<br>(789) | 13.7<br>(175) |
| Drinks less/less frequently | 58.4<br>(921) | 26.9<br>(314) | 14.2<br>(343) | 15.3<br>(497) | 15.2<br>(394) | 42.3<br>(3217) | 14.3<br>(318) | 35.1<br>(730) | 38.9<br>(496) |

For more information about the questions asked in each dataset, and how the change variables were coded, please see Supplementary File 2. Percentages are weighted (except GS). Note:
Analysis for GS, USOC, and ELSA restricted to participants aged 66 and younger. NA= Not Available.

Proportions drinking alcohol on more than four days a week had risen considerably in many studies (e.g. 28% to 52% in USOC) but not all (9% to 10% in MCS), and even where prevalence had risen many participants still reported reducing the frequency of their drinking. In contrast, all studies showed either overall decline or stability in reporting five or more drinks per typical drinking occasion (e.g. 31% to 11% in MCS), though again, a range of 9-31% of participants across all studies also reported consuming increased quantities of alcohol per occasion during the pandemic. When combining frequency and quantity measures, increases in frequency tended to dominate over the decreases in quantity (the young MCS cohort being an exception).

### Pooled Analysis

Figure 2 shows meta-analysis estimates from unadjusted, basic adjusted, and fully adjusted models for smoking, vaping and alcohol consumption during the pandemic. Figure 3 shows pooled estimates from fully adjusted models stratified by sex, education and age. Stratified estimates were largely consistent with the main results, though we highlight some differences below. Full details of the meta-analysis including overall and stratified estimates from each study are available in Supplementary File 3.

**Figure 2:**
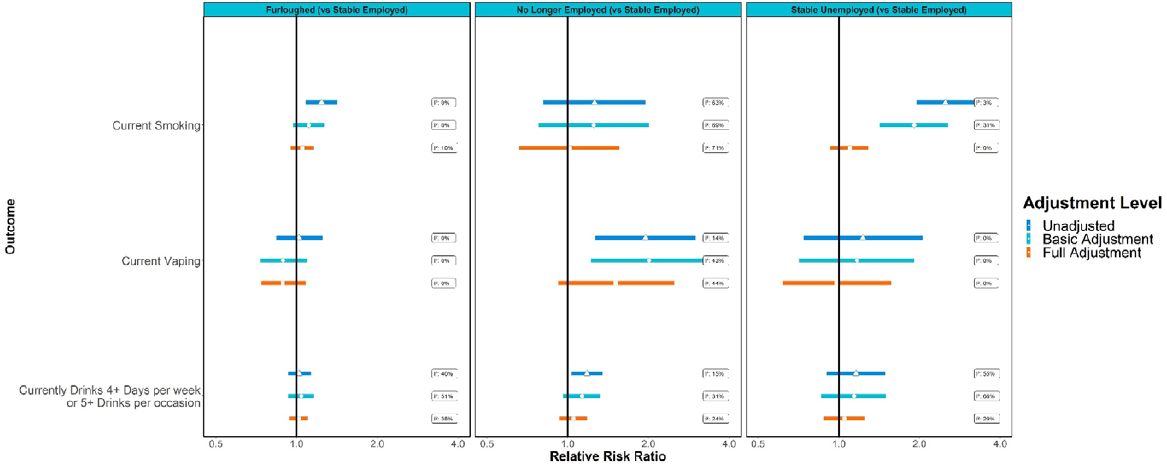
Associations between changes in employment status during the pandemic and health behaviours in pooled analyses across eight UK longitudinal studies. ‘Basic’ adjustment includes age, sex, ethnicity, education, UK nation, and household composition. ‘Full’ adjustment additionally includes pre-pandemic measures of mental health, self-rated health, smoking, vaping and drinking.

**Figure 3:**
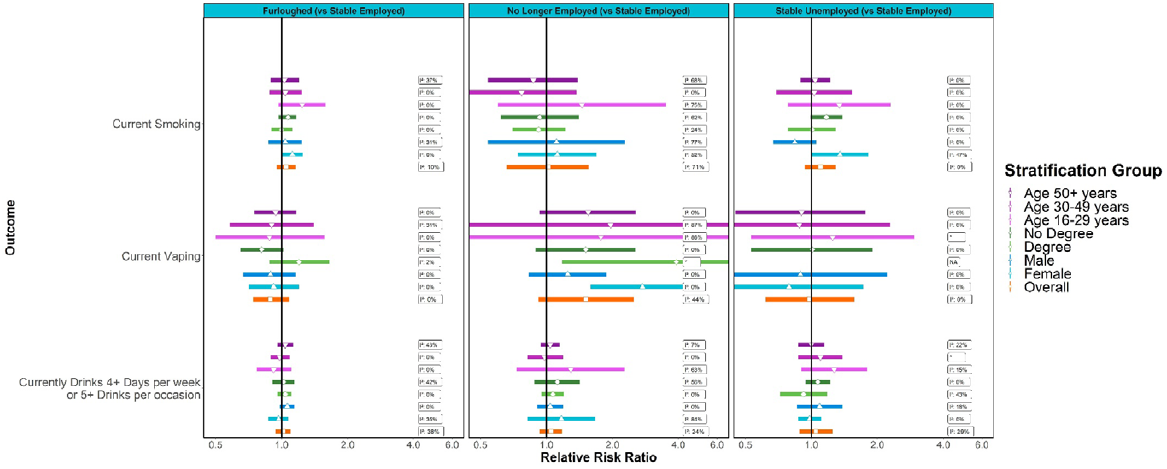
Associations between changes in employment status during the pandemic and health behaviours, stratified by age, sex and educational attainment. *No I^2^ value as only one study was able to provide an estimate. NA=no studies were able to provide reliable estimates.

#### Furlough

Compared to stable employment, unadjusted esimates indicated greater risk of smoking among participants who were furloughed (RR=1.24; 95% CI: 1.08-1.42; I^2^: 0%), but little difference in risk of vaping (RR=1.02; 95% CI: 0.84-1.25; I^2^: 31%) or drinking (RR=1.02; 95% CI: 0.93-1.13; I^2^: 40%). The difference in smoking was attenuated with adjustment for sociodemographic characteristics (ARR=1.11; 95% CI: 0.97-1.27; I^2^: 0%) and further attenuated with adjustment for pre-pandemic mental health, self-rated health and behaviour (ARR=1.05; 95% CI: 0.95-1.16; I^2^: 10%).

In the stratified analyses with full adjustment, furlough was still associated with smoking among women (ARR=1.12; 95% CI: 1.00-1.25; I^2^: 0%), but this estimate did not clearly differ from that among men (ARR=1.04; 95% CI: 0.87-1.24; I^2^: 31%). There was also evidence that furlough was associated with less risk of vaping for those without degree-level education (ARR=0.81; 95% CI: 0.65-1.02; I^2^: 0%), with the association in the opposite direction for those with a degree (ARR=1.20; 95% CI: 0.88-1.65; I^2^: 2%).

#### No Longer Employed

Compared to stable employment, those no longer employed during the pandemic had increased risk of vaping (RR=1.95; 95% CI: 1.27-3.00; I^2^: 14%) and drinking (RR=1.18; 95% CI: 1.03-1.35; I^2^: 15%) and there was some indication of increased risk for smoking (RR=1.26; 95% CI: 0.81-1.95; I^2^: 63%), but confidence intervals were wide and crossed the null. Adjusting for sociodemographic characteristics, mildly attenuated the estimate for drinking (ARR=1.13; 95% CI: 0.96-1.32; I^2^: 31%), while those for vaping and smoking were largely unchanged. No clear differences remained in the overall analyses when we additionally adjusted for pre-pandemic health and behaviour.

In stratified analyses however, even with full adjustment, the association between no longer being employed and risk of vaping was clearly discernible among women (ARR=2.74; 95% CI: 1.59-4.72; I^2^: 0%) but weaker for men (ARR=1.25; 95% CI: 0.83-1.87; I^2^: 0%).

#### Stable Unemployment

Participants in stable unemployment had raised risk of current smoking (RR=2.50; 95% CI: 1.95-3.21; I^2^: 3%) compared to stable employment, and this excess risk was of greater magnitude than that associated with being furloughed or no longer employed. No clear differences were present for vaping or drinking. The difference in risk for smoking was partially attenuated with adjustment for sociodemographic characteristics (ARR=1.91; 95% CI: 1.42-2.56; I^2^: 31%), but more fully attenuated with adjustment for pre-pandemic health and behaviours (ARR=1.10; 95% CI: 0.93-1.29; I^2^: 0%).

In stratified analyses, stable unemployment remained associated with risk of smoking for women (ARR=1.35; 95% CI: 1.00-1.82; I^2^: 47%), even after full adjustment, but not for men (0.84; 95% CI: 0.67-1.05; I^2^: 0%).

#### Changes in Behaviour

Pooled estimates for outcomes indicating changes in smoking, vaping or drinking behaviour are presented in supplementary file 3. Compared to stable employment, and with adjustment for sociodemographic and other pre-pandemic characteristics, no longer being employed and stable unemployment were both associated with smoking less than before the pandemic began, while furlough and no longer being employed were both associated with vaping more than before the pandemic began. The association between furlough and increased vaping was particularly evident among those with degree-level education (ARR: 2.93; 95% CI: 1.80-4.75; I^2^: 0%), rather than those without (ARR: 1.31; 95% CI: 0.79-2.19; I^2^: 51%).

The only overall association with change in drinking behaviour was that participants in stable unemployment were less likely to report increased drinking (ARR: 0.70; 95% CI: 0.53-0.92; I^2^: 28%). However, this association was stronger among those aged under 50 years (e.g. 30-49 year old ARR: 0.38; 95% CI: 0.21-0.70; I^2^: 0%), and weaker for those aged 50 or more (ARR: 0.85; 95% CI: 0.69-1.06; I^2^: 0%). Furlough was associated with increased drinking for men (ARR: 1.33; 95% CI: 1.08-1.64; I^2^: 74%) but not women (ARR: 1.00; 95% CI: 0.90-1.11; I^2^: 34%) while stable unemployment was associated with reduced drinking for those with a degree (ARR: 1.61; 95% CI: 1.05-2.49; I^2^: 39%) but not those without (ARR: 0.83; 95% CI: 0.65-1.06; I^2^: 0%).

## Discussion

We conducted co-ordinated analyses in eight UK longitudinal population surveys and found that, compared to stable employment, furlough was associated with greater risk of smoking during the early stages of the COVID-19 pandemic (April-July 2020). Participants in stable unemployment had a higher magnitude of excess risk for smoking than that associated with furlough, while the magnitude of excess risk for smoking associated with being no longer employed was similar to that for furlough (with wide confidence intervals). Furlough was not associated with excess risk of vaping or drinking, compared to stable employment, while no longer being employed was associated with higher risk for both outcomes. All of these differences were largely explained when accounting for sociodemographic characteristics and pre-pandemic health (mental and physical) and behaviour, though this did differ by gender. Among women but not men, furlough and stable unemployment remained associated with risk of smoking (the magnitude of excess risk was greater for stable unemployment than that for furlough), and no longer being employed remained associated with vaping, even after adjusting for pre-pandemic characteristics.

Previous studies have indicated overall slight declines in smoking and vaping during the pandemic (10), with evidence of increases in smoking cessation attempts and rates of successful quitting (12), though there has been little evidence as yet on the role of employment disruptions during the pandemic. Job loss has been associated with increased smoking in previous studies (15, 16), but it has not been clear whether ceasing work as part of a furlough scheme or in a pandemic would have the same effect and could have contributed to changes in drinking behaviour. In our data from the initial months of the pandemic in the UK, once we took potential confounders including pre-pandemic behaviours into account we found no convincing evidence of an effect of furlough on these health behaviours. Furlough aside, we also did not find the usual associations between unemployment (new or stable) and smoking (except for the association between stable unemployment and smoking among women). Indeed, supplementary analyses of behaviour change indicated that unemployment was if anything associated with reductions in smoking during the pandemic.

Previous research has detailed increases in frequency of drinking and of binge drinking during the pandemic (10, 13), though there has also been evidence of polarisation in drinking patterns, with some of the younger cohorts studied here reducing their drinking (46). Unemployment has been associated with heavier drinking in previous studies (16, 17), though as discussed above, it has not been clear whether this would remain true in a pandemic, or whether similar effects should be expected for furlough. Initial surveys of small-scale and self-selected samples have indicated an association between furlough and increased drinking (14, 18). We found weak support for this in large scale nationally representative surveys, but only among men and only when specifically analysing self-reported change in drinking. There was no clear association between furlough and alcohol consumption during the pandemic in our main analyses, either before or after adjustment for pre-pandemic characteristics (including pre-pandemic drinking). This discrepancy could be because: associations with change outcomes can include effects of employment on pre-pandemic drinking; or because analyses of change were sensitive to relatively minor changes in behaviour above or below the threshold used to identify heavy drinking in the main analyses (which may have less overall relevance for health). The measurement difference may be especially important, as one of the previous studies only found an association with furlough for heavy episodic drinking (14), rather than for frequency or quantity measures similar to those derived here.

While combining analyses from several UK prospective studies represents a clear contribution to understanding the potential impact of furlough, there are limitations that should be taken into account. Firstly, we were not able to achieve full harmonisation of measures across studies, for example, a number of studies had only asked questions on recent pre-pandemic smoking of those who were smoking during the pandemic, meaning smoking cessation during the pandemic was unobserved in those studies. This means the analyses of change in smoking for these studies focused only on reductions in cigarettes smoked, rather than outright cessation. The main analyses will be less affected, though there may have been some residual confounding from participants who had smoked but given up before being surveyed during the pandemic. Focusing on maximising comparability of measures across studies also limited our scope to explore more varied definitions with respect to frequency, quantity or other aspects of use, such as binge drinking, or concurrence of smoking and vaping behaviour. Our findings focus on the initial stages of the pandemic (April-July 2020) and observed relationships could change with the duration of lockdown or furlough, and subsequent changes to restrictions and the furlough scheme. Further research is needed to explore potential heterogeneity over the course of the pandemic, within workers who retained employment, and between different measurement definitions/thresholds.

As with most observational studies, unobserved confounding could have affected our estimates. For example, we did not adjust for occupational class (since it was unobservable for those who were not employed), and there may have been differences between participants whose jobs were retained, versus those who experienced furlough or job loss. Our fully adjusted models account for differences in some key pre-pandemic characteristics among employment groups, but it is possible that our results reflect other traits of these employment groups, for example, how workers in different industries or occupational classes were responding to the pandemic, rather than being effects of furlough specifically. Furthermore, despite being embedded in long standing cohorts, survey responses during the pandemic were selective, and while weighting was employed to correct for this, bias due to selective non-response cannot be excluded (47). Also, while adjustment for pre-pandemic characteristics was important, it may also have introduced bias in estimates if there were unobserved determinants of both pre-pandemic characteristics and behaviour during the pandemic (48). We found participation in the furlough scheme to be considerably more common during the initial stages of the pandemic than being no longer employed or in stable unemployment, which meant that estimates for the latter groups were based on small numbers with considerable heterogeneity and imprecision in estimates, especially in stratified analyses, or for rarer outcomes like vaping, or change in smoking. Nevertheless, confidence interval aside, even the estimated magnitude of the associations for no longer being employed and stable unemployment were close to the null after adjusting for pre-pandemic characteristics (the association between no longer being employed and risk of vaping was the clearest exception to this pattern). Furthermore, our findings need to be interpreted in the context of the pandemic and associations for moving out of employment in particular are likely to differ in comparison to when the labour market is functioning under more typical circumstances.

Since new unemployment has previously been associated with increases in smoking and drinking (15–17), there was some potential for a furlough scheme to have adverse impacts on smoking and drinking behaviour, though the preservation of income and recognisably temporary nature of furlough may have softened impacts usually associated with ceasing work. While furlough was associated with higher prevalence of smoking, we found little evidence to support any adverse impacts after adjusting for pre-pandemic characteristics. After such adjustment, we did find that furlough was associated with increased alcohol consumption among men, and with higher risk of smoking among women, but these associations were weak enough to be plausibly explained by unmeasured confounding. The association with increased drinking for men was not consistently apparent across all analyses, as would be expected if it were a strong and robust causal relationship, and there was not clear evidence that the association with smoking for women was actually stronger than that among men: the overall analyses did not show a clear relationship. There was also an indication that furlough was associated with increased vaping, but some or all of this may represent moves towards smoking cessation. Thus, furlough does not appear to have had any clear adverse impacts on smoking, vaping or drinking behaviour.

A qualitative study on smoking during the pandemic identified stresses associated with confinement, curtailment of social routines, removal of barriers and distractions, and feelings of boredom as mechanisms that could contribute to increased smoking (49). Many of these could be relevant for either furlough or unemployment and could affect drinking behaviour too. Nevertheless, associations might not be apparent if these mechanisms were also relevant to people in stable employment who would still have had their life and working patterns disrupted and may, for example, have had to start working from home or make other adaptations. The ubiquity of pandemic related disruptions may help explain the lack of associations observed overall, with clear differences only emerging in sub-groups, or additional analyses of behaviour change.

With respect to vaping, the clearest association observed was for workers who were no longer employed during the pandemic. We are inclined to interpret this cautiously, since numbers in this group were small and vaping was rare, but positively since most vapers were or had been smokers, and this group also exhibited a greater likelihood of decreasing their cigarette use. While not what our study was designed to address, this is consistent with e-cigarettes being used as an aid to reducing or quitting smoking (50, 51), especially among a disadvantaged group.

### Conclusion

Alongside the beneficial role of furlough in mitigating adverse economic impacts of the pandemic, there is little evidence of it having any detrimental impacts on smoking, vaping and drinking behaviours relative to remaining in employment. Indeed, neither no longer being employed nor stable unemployment were clearly associated with these behaviours after accounting for pre-pandemic characteristics either. Thus, employment status disruption does not appear to have been a clear driver of smoking, vaping or drinking behaviours during the early stages of the pandemic.

## Data Availability

All datasets included in this analysis have established data sharing processes, and for most included studies the anonymised datasets with corresponding documentation can be downloaded for use by researchers from the UK Data Service. We have detailed the processes for each dataset in Supplementary File 1.

## List of abbreviations

ARR: Adjusted Risk Ratio
ALSPAC-G1: Avon Longitudinal Study of Parents and Children
ALSPAC-G0: Parents of ALSPAC-G1
BCS70: 1970 British Cohort Study
CI: Confidence interval
CJRS: Coronavirus Job Retention Scheme
ELSA: English Longitudinal Study of Ageing.
GS: Generation Scotland: the Scottish Family Health Study
MCS: Millennium Cohort Study
NCDS: 1958 National Child Development Study
NS: Next Steps (formerly the Longitudinal Study of Young People in England)
UK: United Kingdom
USOC: Understanding Society

## Additional Files

Supplementary File 1: Study Description

Supplementary File 2: Variable Coding

Supplementary File 3: Meta-Analysis

Supplementary File 4: Figures

Supplementary File 5: Tables

Supplementary File 6: Supplementary Analyses of Drinking

## Declarations

### Ethics approval and consent to participate

We have detailed the ethical approval for each study in Supplementary File 1.

### Author Contribution Statement

Conceptualised the study and design: Green, Maddock, Di Gessa, Wielgoszewska, Parsons, Ploubidis, Katikireddi

Designed the methodology: Green, Maddock, Di Gessa, Wielgoszewska, Parsons, Ploubidis, Katikireddi, Griffith and Silverwood

Conducted the formal analysis: Green, Maddock, Di Gessa, Wielgoszewska, Parsons Croft, Stevenson, Huggins, Booth and Griffith

Data curation: Green, Maddock, Di Gessa, Stevenson, Huggins, Griffith, Chaturvedi, Howe and Fitzsimons

Wrote the original draft of the manuscript: Green, Maddock, Di Gessa, Wielgoszewska, Parsons.

Data visualisation: Green and Di Gessa.

All authors contributed to critical revision of the manuscript.

Supervision: Ploubidis and Katikireddi.

Funding acquired: Patalay, Katikireddi, Ploubidis, Silverwood, and Chaturvedi.

## Acknowledgements

The contributing studies have been made possible because of the tireless dedication, commitment and enthusiasm of the many people who have taken part. We would like to thank the participants and the numerous team members involved in the studies including interviewers, technicians, researchers, administrators, managers, health professionals and volunteers. We are additionally grateful to our funders for their financial input and support in making this research happen.

GS: Drew Altschul, Chloe Fawns-Ritchie, Archie Campbell, Robin Flaig.

ALSPAC: Daniel J Smith, Nicholas J Timpson, Kate Northstone

Understanding Society: Michaela Benzeval

MCS, NS, BCS70, NCDS: Colleagues in survey, data and cohort maintenance teams

