## Supplementary File 1 for "The UK COVID-19 furlough scheme and associations with smoking, alcohol consumption and vaping: evidence from 8 UK longitudinal population surveys"

### **S1.1 Description of Studies**

| **Study Population** | **Design, Sample Frame and Weighting** | **2020 Age range included in years** | **Most recent pre-pandemic survey** | **Details of 2020 covid surveys used**  **(response rate)** | **Key Citations and Reference**  **(see S**1.2 Study **Reference List)** | **Analytical N** |
| --- | --- | --- | --- | --- | --- | --- |
| *Age Homogenous Cohorts* | |  |  |  |  |  |
| MCS: Millennium Cohort Study | A nationally representative cohort of UK children born between Sept 2000 and Jan 2002 with regular follow-up surveys from birth. Weighted for sampling design, pre-pandemic attrition and non-response to COVID surveys. | 18-20 | 2018-2019 | May (26.6%) | 1, 2 | 2057 |
| ALSPAC (G1): Avon Longitudinal Study of Parents and Children- Generation 1 (original young people) | Cohort of children born in the South-West of England between April 1991 and Dec 1992, with regular follow-up surveys from birth.  Weighted for pre-pandemic attrition and non-response to COVID surveys. | 27-29 | 2017-2018 | June (17.4%) | 3, 4 | 1275 |
| NS: Next Steps, formerly known as Longitudinal Study of Young People in England | A nationally representative sample recruited via secondary schools in England at around age 13 with regular follow-up surveys thereafter. Weighted for sampling design, pre-pandemic attrition and non-response to COVID surveys. | 29-31 | 2015 | May (20.3%) | 2, 5 | 1579 |
| BCS70: British Cohort Study 1970 | A nationally representative cohort of all children born in Great Britain (i.e. England, Wales & Scotland) in one week in 1970, with regular follow-up surveys from birth. Weighted for pre-pandemic attrition and non-response to COVID surveys. | 50 | 2016 | May (40.4%) | 2, 6 | 3151 |
| NCDS: National Child Development Study | A nationally representative cohort of all children born in Great Britain (i.e. England, Wales & Scotland) in one week in 1958, with regular follow-up surveys from birth. Weighted for pre-pandemic attrition and non-response to COVID surveys. | 62 | 2013 | May (57.9%) | 2, 7 | 4358 |
| *Age Heterogeneous Studies* | |  |  |  |  |  |
| USOC: Understanding Society: the UK Household Longitudinal Survey | A nationally representative longitudinal household panel study, based on a clustered-stratified probability sample of UK households, with all adults aged 16+ in chosen households surveyed annually. Weighted for sampling design, pre-pandemic attrition, non-response to COVID surveys, and outcome non-response within COVID surveys. | 17-66 | 2018-2019 | April (40.3%) | 8, 9 | 8328 |
| ELSA: English Longitudinal Study of Aging | A nationally representative population study of individuals aged 50+ living in England, with biennial surveys and periodic refreshing of the sample to maintain representativeness. Weighted for sampling design, pre-pandemic attrition, and non-response to COVID surveys. | 52-66 | 2018-2019 | Jun-July (75%) | 10, 11 | 2417 |
| GS: Generation Scotland: the Scottish Family Health Study | A family-structured, population-based Scottish cohort, with participants aged 18-99 recruited between 2006-2011. No weights were available. | 27-66 | 2006-2011 | April-Jun (21.6%) | 12, 13 | 2604 |
| ALSPAC(G0): Avon Longitudinal Study of Parents and Children- Generation 0 (original parents) | Parents of the ALSPAC(G1) cohort described above, treated as a separate age-heterogenous study population. | 44-66 | 2011-2013 | June (12.2%) | 4, 14 | 2072 |

**S1.2 Study Reference List**

1. Joshi H, Fitzsimons E. The UK Millennium Cohort Study: the making of a multi-purpose resource for social science and policy in the UK. Longitudinal and Life Course Studies. 2016;7(4):409-30.
2. Brown, M., Goodman, A., Peters, A., Ploubidis, G.B., Sanchez, A., Silverwood, R., Smith, K. (2021) COVID-19 Survey in Five National Longitudinal Studies: Waves 1, 2 and 3 User Guide (Version 3). London: UCL Centre for Longitudinal Studies and MRC Unit for Lifelong Health and Ageing.
3. Boyd A, Golding J, Macleod J, Lawlor DA, Fraser A, Henderson J, et al. Cohort Profile: the 'children of the 90s'--the index offspring of the Avon Longitudinal Study of Parents and Children. Int J Epidemiol. 2013;42(1):111-27.
4. Northstone K, Smith D, Bowring C et al. The Avon Longitudinal Study of Parents and Children - A resource for COVID-19 research: Questionnaire data capture May-July 2020. Wellcome Open Res 2020, 5:210 (<https://doi.org/10.12688/wellcomeopenres.16225.1>).
5. Calderwood L, Sanchez C. Next Steps (formerly known as the Longitudinal Study of Young People in England). Journal of Open Health Data. 2016;4.
6. Elliott J, Shepherd P. Cohort profile: 1970 British Birth Cohort (BCS70). Int J Epidemiol. 2006;35(4):836-43.
7. Power C, Elliott J. Cohort profile: 1958 British birth cohort (National Child Development Study). Int J Epidemiol. 2006;35(1):34-41.
8. University of Essex, Institute for Social and Economic Research, NatCen Social Research, Kantar Public. Understanding Society: Waves 1-10, 2009-2017 and Harmonised BHPS: Waves 1-18, 1991-2009. 13th Edition ed: UK Data Service; 2020.
9. Institute for Social and Economic Research (2021) Understanding Society COVID-19 User Guide. Version 9.0, July 2021. Colchester: University of Essex.
10. Steptoe A, Breeze E, Banks J, Nazroo J. Cohort profile: the English longitudinal study of ageing. Int J Epidemiol. 2013;42(6):1640-8.
11. Steptoe, A., Addario, G., Banks, J., Batty, G. David, Coughlin, K., Crawford, R., Dangerfield, P., Marmot, M., Nazroo, J., Oldfield, Z., Pacchiotti, B., Steel, N., Wood, M., Zaninotto, P. (2021). English Longitudinal Study of Ageing COVID-19 Study, Waves 1-2, 2020. [data collection]. 2nd Edition. UK Data Service. SN: 8688, <http://doi.org/10.5255/UKDA-SN-8688-2>.
12. Smith BH, Campbell A, Linksted P, Fitzpatrick B, Jackson C, Kerr SM, et al. Cohort Profile: Generation Scotland: Scottish Family Health Study (GS:SFHS). The study, its participants and their potential for genetic research on health and illness. Int J Epidemiol. 2013;42(3):689-700.
13. Fawns-Ritchie C, Altschul DM, Campbell A et al. CovidLife: a resource to understand mental health, well-being and behaviour during the COVID-19 pandemic in the UK. Wellcome Open Res 2021, 6:176 (<https://doi.org/10.12688/wellcomeopenres.16987.1>).
14. Fraser A, Macdonald-Wallis C, Tilling K, Boyd A, Golding J, Davey Smith G, Henderson J, Macleod J, Molloy L, Ness A, Ring S, Nelson SM, Lawlor DA. Cohort Profile: The Avon Longitudinal Study of Parents and Children: ALSPAC mothers cohort. International Journal of Epidemiology 2013; 42:97-110.

### **S1.3 Ethics and data access statements for each study**

The most recent sweeps of the  **NCDS, BCS70**, **Next Steps** and **MCS** have all been granted ethical approval by the National Health Service (NHS) Research Ethics Committee and all participants have given informed consent. Data for NCDS (SN 6137), BCS70 (SN 8547), Next Steps (SN 5545), MCS (SN 8682) and all four COVID-19 surveys (SN 8658) are available through the UK Data Service. NSHD data are available on request to the NSHD Data Sharing Committee. Interested researchers can apply to access the NSHD data via a standard application procedure. Data requests should be submitted to; further details can be found at <http://www.nshd.mrc.ac.uk/data.aspx>. doi:10.5522/NSHD/Q101; doi:10.5522/NSHD/Q10.

Ethical approval was obtained from the **ALSPAC** Ethics and Law Committee and the Local Research Ethics Committees. The study website contains details of all the data that is available through a fully searchable data dictionary and variable search tool: <http://www.bristol.ac.uk/alspac/researchers/our-data>. ALSPAC data is available to researchers through an online proposal system. Information regarding access can be found on the ALSPAC website (<http://www.bristol.ac.uk/media-library/sites/alspac/documents/researchers/data-access/ALSPAC_Access_Policy.pdf>).

All wave of **TwinsUK** have received ethical approval associated with TwinsUK Biobank (19/NW/0187), TwinsUK (EC04/015) or Healthy Ageing Twin Study (H.A.T.S) (07/H0802/84) studies from NHS Research Ethics Committees at the Department of Twin Research and Genetic Epidemiology, King’s College London. The TwinsUK Resource Executive Committee (TREC) oversees management, data sharing and collaborations involving the TwinsUK registry (for further details see <https://twinsuk.ac.uk/resources-for-researchers/access-our-data/>).

The University of Essex Ethics Committee has approved all data collection for the **Understanding Society** main study and COVID-19 waves. No additional ethical approval was necessary for this secondary data analysis. All data are available through the UK Data Service (SN 6614 and SN 8644).

Waves 1-9 of **ELSA** were approved through the National Research Ethics Service, while the COVID-19 Sub-study was approved by the UCL Research Ethics Committee. All participants provided informed consent. All data are available through the UK Data Service (SN 8688 and 5050).

**Generation Scotland** obtained ethical approval from the East of Scotland Committee on Medical Research Ethics (on behalf of the National Health Service). Reference number 20/ES/0021. Access to data is approved by the Generation Scotland Access Committee. See <https://www.ed.ac.uk/generation-scotland/for-researchers/access> or for further details.
