## Supplementary figures and images for "The UK COVID-19 furlough scheme and associations with smoking, alcohol consumption and vaping: evidence from 8 UK longitudinal population surveys"

### Supplementary File 4

**Supplementary Figure S1: Causal pathways blocked under differing levels of adjustment**

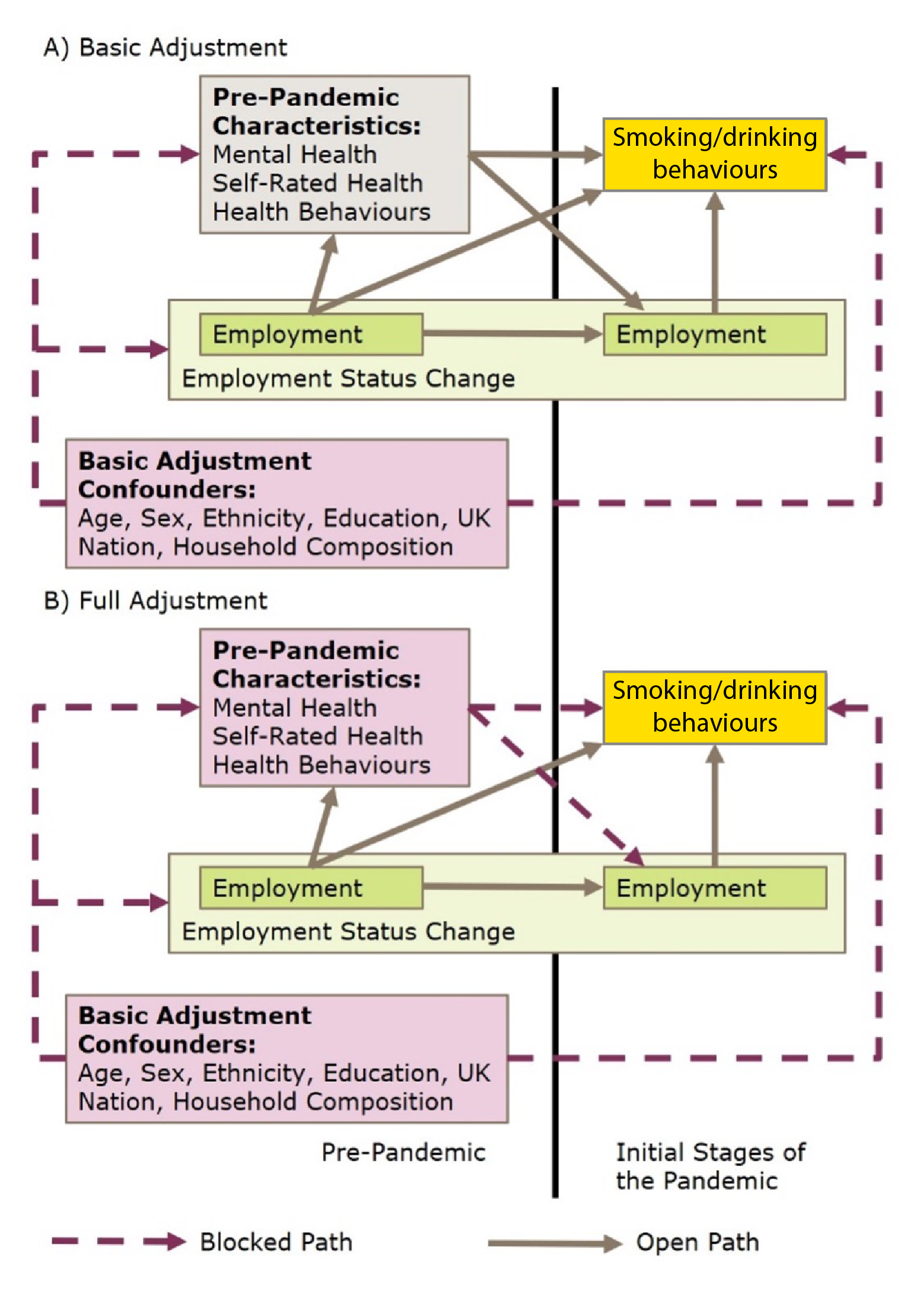
