## Supplementary File 5 for "The UK COVID-19 furlough scheme and associations with smoking, alcohol consumption and vaping: evidence from 8 UK longitudinal population surveys"

**Supplementary Table S5.1: Descriptive Characteristics by Study**

|  | **MCS** | **NS** | **BCS** | **NCDS** | **GS** | **USOC** | **ELSA** | **ALSPAC-G0** | **ALSPAC-G1** |
| --- | --- | --- | --- | --- | --- | --- | --- | --- | --- |
| Total N | 2057 | 1579 | 3151 | 4358 | 2604 | 8,328 | 2,417 | 2072 | 1275 |
| Age/Age range | 18-20 | 29-31 | 50 | 62 | 27-66 | 17-66 | 52-66 | 50-65 | 27-29 |
|  | % | % | % | % | % | % | % | % | % |
| *Gender* |  |  |  |  |  |  |  |  |  |
| Male | 49.6  (46.3-52.9) | 44.1  (40.0-48.4) | 48.0  (46.3-49.8) | 49.4  (47.9-50.9) | 33.5  (31.7-35.3) | 47.9  (46.5-49.2) | 48.0  (45.6-50.5) | 22.8  (20.5-25.4) | 29.0  (25.6-32.7) |
| Female | 50.4  (47.1-53.7) | 55.9  (51.6-60.0) | 52.0  (50.2-53.7) | 50.6  (49.1-52.1) | 66.5  (64.7-68.3) | 52.1  (50.8-53.5) | 52.0  (49.5-54.4) | 77.2  (74.6-79.5) | 71.0  (67.3-74.4) |
| *Ethnicity* |  |  |  |  |  |  |  |  |  |
| White | 88.3  (84.9-91.1) | 87.6  (85.0-89.8) | NA | NA | 99.2  (98.8-99.5) | 89.6  (88.2-90.8) | 90.2  (88.2-91.8) | 98.8  (98.0-99.2) | 96.5  (94.3-97.9) |
| Non-White Ethnic Minority | 11.7  (8.9-15.1) | 12.4  (10.2-15.0) | NA | NA | 0.8  (0.5-1.2) | 10.4  (9.2-11.8) | 9.8  (8.2-11.8) | 1.2  (0.7-2.0) | 3.5  (2.1-5.8) |
| *Education* |  |  |  |  |  |  |  |  |  |
| Degree | 39.4  (34.9-44.0) | 45.3  (41.3-49.3) | 39.7  (38.0-41.4) | 35.4  (33.9-36.8) | 52.1  (50.2-54.0) | 39.6  (38.1-41.0) | 25.0  (23.0-27.1) | 20.8  (19.0-22.8) | 30.6  (27.1-34.3) |
| Less than degree | 60.6  (56.0-65.1) | 54.7  (50.7-58.7) | 60.3  (58.6-62.0) | 64.6  (63.2-66.1) | 47.9  (46.0-49.8) | 60.4  (59.0-61.9) | 75.0  (73.0-77.0) | 79.2  (77.2-81.0) | 69.4  (65.7-72.9) |
| *Household Composition* |  |  |  |  |  |  |  |  |  |
| Single, no children/ Alone | 2.3  (1.2-4.2) | 10.9  (8.8-13.4) | 14.2  (13.0-15.4) | 24.0  (22.7-25.3) | 13.0  (11.8-14.4) | 39.9  (38.3-41.6) | 18.3  (16.3-20.4) | 6.8  (5.4-8.6) | 8.3  (6.3-10.9) |
| Couple, no children/ Living only with partner | 2.5  (1.8-3.6) | 31.6  (28.0=35.4) | 18.2  (16.9-19.6) | 46.9  (45.4-48.4) | 39.4  (37.6-41.3) | 30.9  (29.7-32.2) | 38.4  (36.2-40.8) | 33.3  (30.8-36.0) | 46.7  (42.8-50.7) |
| Single, with children | NA | NA | NA | NA | 4.8  (4.0-5.6) | 4.0  (3.4-4.8) | 8.7  (7.3-10.5) | NA | NA |
| Couple, with children | 4.0  (2.9-5.5) | 34.6  (30.7-38.8) | 55.4  (53.7-57.2) | 22.5  (21.3-23.8) | 35.4  (33.6-37.3) | 25.1  (23.7-26.5) | 30.8  (28.6-33.2) | 20.8  (18.7-23.1) | NA |
| Others | 91.2  (89.0-93.1) | 23.0  (19.6-26.7) | 12.2  (11.1-13.4) | 6.6  (5.9-7.4) | 7.4  (6.4-8.4) | NA | 3.7  (3.0-4.7) | 33.7  (31.1-36.5) | 34.3  (30.7-37.2) |
| *UK Nation* |  |  |  |  |  |  |  |  |  |
| England | 83.6  (80.8-86.0) | 96.7  (94.7-97.9) | 86.4  (85.2-87.6) | 85.2  (84.1-86.2) | 0.5  (0.3-0.9) | 86.5  (85.4-87.6) | 100.0 | NA | NA |
| Wales | 5.5  (4.3-7.0) | 1.3  (0.3-3.6) | 5.2  (4.5-6.0) | 4.4  (3.8-5.0) | NA | 4.0  (3.5-4.5) | NA | NA | NA |
| Scotland | 8.3  (6.7-10.2) | 0.7  (0.2-1.6) | 7.4  (6.6-8.4) | 8.3  (7.5-9.2) | 99.4  (99.1-99.7) | 7.3  (6.4-8.3) | NA | NA | NA |
| Northern Ireland | 2.0  (1.4-2.7) | 0.1  (0.0-0.4) | 0.2  (0.0-0.4) | 0.3  (0.2-0.6) | NA | 2.2  (1.9-2.5) | NA | NA | NA |
| Other (Channel Islands / Isle of Man | 0.7  (0.1-2.4) | 1.2  (0.8-1.9) | 0.8  (0.5-1.2) | 1.7  (1.4-2.2) | 0.1  (0.0-0.3) | NA | NA | NA | NA |
| *Pre-Pandemic Mental Health* |  |  |  |  |  |  |  |  |  |
| Low to no symptoms | 81.8  (78.8-84.4) | 74.6  (70.9-78.1) | 81.0  (79.5-82.3) | 85.4  (84.3-86.4) | 87.3  (85.9-88.5) | 77.7  (76.4-79.0) | 86.3  (84.4-88.0) | 81.1  (78.6-83.3) | 77.1  (73.5-80.3) |
| High symptoms | 18.2  (15.6-21.2) | 25.4  (21.9-29.1) | 19.0  (17.7-20.5) | 14.6  (13.6-15.7) | 12.7  (11.5-14.1) | 22.3  (21.0-23.6) | 13.7  (12.0-15.6) | 18.9  (16.7-21.4) | 22.9  (19.7-26.5) |
| *Pre-Pandemic Self-Rated Health* |  |  |  |  |  |  |  |  |  |
| Excellent-Good | 85.9  (83.2-88.3) | 88.1  (84.9-90.8) | 85.2  (83.9-86.4) | 81.6  (80.4-82.7) | NA | 80.3  (78.9-81.6) | 77.1  (74.7-79.3) | NA | NA |
| Fair-Poor | 14.1  (11.7-16.8) | 11.9  (9.2-15.1) | 14.8  (13.6-16.1) | 18.4  (17.3-19.6) | NA | 19.7  (18.4-21.1) | 22.9  (20.7-25.3) | NA | NA |

Percentages are weighted (except GS). Note: Analysis for GS, USOC, and ELSA restricted to participants aged 66 and younger. NA= Not Available

**Supplementary Table S5.2: Employment Change by Gender, Education and Age**

|  | **MCS** | **NS** | **BCS** | **NCDS** | **GS** | **USOC** | **ELSA** | **ALSPAC-G0** | **ALSPAC-G1** |
| --- | --- | --- | --- | --- | --- | --- | --- | --- | --- |
| Total N | 2057 | 1579 | 3151 | 4358 | 2604 | 8,328 | 2,417 | 2072 | 1275 |
| Age/Age range | 18-20 | 29-31 | 50 | 62 | 27-66 | 17-66 | 52-66 | 50-66 | 27-29 |
|  | % | % | % | % | % | % | % | % | % |
| **Stable Employed** | 13.4  (11.1-16.1) | 59.6  (55.3-63.8) | 61.5  (59.8-63.2) | 32.8  (31.4-34.2) | 62.7  (60.8-64.5) | 59.0  (57.4-60.5) | 51.1  (48.6-53.6) | 54.7  (51.8-57.5) | 69.1  (66.3-71.7) |
| Male | 13.6  (10.6-17.4) | 68.8  (62.2-74.7) | 65.3  (62.7-67.9) | 35.2  (33.1-37.3) | 65.6  (62.4-68.7) | 60.4  (58.0-62.8) | 55.3  (51.4-59.1) | 53.7  (47.5-59.8) | 71.0  (66.2-75.4) |
| Female | 13.1  (9.9-17.0) | 52.3  (47.2-57.4) | 58.0  (55.7-60.2) | 30.5  (28.7-32.4) | 61.2  (58.9-63.5) | 57.6  (55.7-59.5) | 47.2  (44.1-50.3) | 55.0  (51.8-58.1) | 68.2  (64.8-71.5) |
| Degree | 9.4  (6.1-14.2) | 69.8  (64.3-74.7) | 70.0  (67.7-72.2) | 34.2  (32.2-36.3) | 67.4  (64.8-69.8) | 70.6  (68.8-72.4) | 54.9  (50.5-59.3) | 49.7  (45.0-54.3) | 73.2  (68.0-77.9) |
| No Degree | 16.0  (13.3-19.2) | 51.2  (45.3-57.0) | 55.9  (53.4-58.3) | 32.1  (30.2-34.0) | 57.6  (54.8-60.3) | 51.3  (49.1-53.5) | 49.8  (46.9-52.8) | 56.0  (52.6-59.3) | 67.4  (64.1-70.6) |
| Age 16-29 | 13.4  (11.1-16.1) |  |  |  | 73.0  (57.0-84.6) | 49.2  (45.2-53.1) |  |  | 69.1  (66.3-71.7) |
| Age 30-49 |  | 59.6  (55.3-63.8) |  |  | 78.2  (75.2-80.9) | 69.3  (66.9-71.6) |  |  |  |
| Age 50-66 |  |  | 61.5  (59.8-63.2) | 32.8  (31.4-34.2) | 55.4  (53.1-57.7) | 54.9  (53.0-56.7) | 51.1  (48.6-53.6) | 54.7  (51.8-57.5) |  |
| **Furloughed** | 16.6  (13.8-19.7) | 25.6  (22.2-29.5) | 24.8  (23.3-26.4) | 21.6  (20.4-22.9) | 8.4  (7.4-9.5) | 14.9  (13.8-16.1) | 13.4  (11.9-15.2) | 12.8  (10.9-15.0) | 16.8  (14.7-19.1) |
| Male | 12.9  (9.2-17.8) | 23.0  (17.7-29.3) | 24.8  (22.5-27.2) | 25.6  (23.7-27.5) | 10.1  (8.3-12.3) | 15.8  (14.1-17.7) | 12.8  (10.4-15.5) | 13.5  (9.4-19.0) | 14.5  (11.1-18.6) |
| Female | 19.8  (16.4-23.6) | 27.7  (23.1-32.8) | 24.9  (23.0-26.9) | 13.8  (12.4-19.4) | 7.6  (5.4-8.9) | 14.1  (12.9-15.5) | 14.1  (12.1-16.4) | 12.8  (10.9-15.0) | 17.8  (15.2-20.7) |
| Degree | 10.7  (8.1-13.9) | 19.0  (14.8-24.1) | 20.4  (18.5-22.5) | 13.8  (12.4-15.4) | 5.1  (4.0-6.4) | 10.6  (9.4-11.9) | 11.5  (9.0-14.6) | 7.9  (5.6-10.9) | 15.9  (12.1-20.6) |
| No Degree | 20.4  (16.6-24.7) | 31.1  (25.9-36.8) | 27.7  (25.6-30.0) | 25.9  (24.2-27.8) | 12.0  (10.3-14.0) | 17.8  (16.2-19.5) | 14.1  (12.2-16.2) | 14.1  (11.8-16.8) | 17.2  (14.7-19.9) |
| Age 16-29 | 16.6  (13.8-19.7) |  |  |  | 2.7  (0.5-13.8) | 17.9  (15.2-20.8) |  |  | 16.8  (14.7-19.1) |
| Age 30-49 |  | 25.6  (22.2-29.5) |  |  | 10.5  (8.5-12.8) | 16.0  (14.2-17.9) |  |  |  |
| Age 50-66 |  |  | 24.8  (23.3-26.4) | 21.6  (20.4-22.9) | 7.6  (6.4-8.9) | 11.8  (10.6-13.0) | 13.4  (11.9-15.2) | 12.8  (10.9-15.0) |  |

|  | **MCS** | **NS** | **BCS** | **NCDS** | **GS** | **USOC** | **ELSA** | **ALSPAC-G0** | **ALSPAC-G1** |
| --- | --- | --- | --- | --- | --- | --- | --- | --- | --- |
| **No Longer Employed** | 3.6  (2.6-5.0) | 2.6  (1.7-3.9) | 1.9  (1.5-2.5) | 2.8  (2.4-3.4) | 3.3 (2.7-4.1) | 3.5  (2.8-4.2) | 2.0  (1.4-2.7) | 7.0  (5.8-8.5) | 5.7  (4.4-7.2) |
| Male | 4.3  (2.6-6.9) | 1.5  (0.8-2.8) | 2.0  (1.3-2.9) | 3.0  (2.3-3.8) | 3.4  (2.4-4.9) | 3.2  (2.3-4.4) | 2.2  (1.3-3.6) | 7.1  (5.7-8.8) | 4.4  (2.8-6.8) |
| Female | 2.9  (2.0-4.0) | 3.4  (2.0-5.6) | 1.9  (1.4-2.6) | 2.7  (2.1-3.4) | 3.3  (2.5-4.2) | 3.7  (2.9-4.8) | 1.8  (1.2-2.6) | 6.6  (4.5-9.7) | 6.8  (6.5-7.1) |
| Degree | 4.5  (3.1-6.6) | 3.2  (1.7-5.7) | 1.8  (1.3-2.6) | 2.8  (2.1-3.6) | 3.3  (2.5-4.4) | 3.0  (2.4-3.8) | 1.5  (0.9-2.6) | 10.2  (7.8-13.3) | 5.2  (3.3-8.2) |
| No Degree | 3.0  (1.8-5.0) | 2.1  (128-3.6) | 2.0  (1.4-2.8) | 2.8  (2.2-3.6) | 3.4  (2.5-4.5) | 3.8  (2.8-5.0) | 2.1  (1.5-3.1) | 6.1  (4.8-7.9) | 5.8  (4.3-7.8) |
| Age 16-29 | 3.6  (2.6-5.0) |  |  |  |  | 5.6  (3.9-7.9) |  |  | 5.7  (4.4-7.2) |
| Age 30-49 |  | 2.6  (1.7-3.9) | 1.9  (1.5-2.5) | 2.8  (2.4-3.4) | 2.7  (1.8-4.1) | 2.6  (1.7-4.0) |  |  |  |
| Age 50-66 |  |  |  |  | 3.7  (2.9-4.7) | 2.9  (2.4-3.5) | 2.0  (1.4-2.7) | 7.0  (5.8-8.5) |  |
| **Stable Unemployed** | 5.8  (3.7-8.8) | 1.8  (1.0-3.1) | 0.9  (0.6-1.3) | 2.1  (1.8-2.6) | 0.5  (0.3-0.8) | 2.4  (1.9-3.0) | 3.1  (2.2-4.4) | 8.3  (6.9-9.9) | 2.9  (2.2-3.8) |
| Male | 8.4  (4.6-14.6) | 1.4  (0.5-3.2) | 1.0  (0.6-1.8) | 2.7  (2.0-3.5) | 0.8  (0.4-1.6) | 2.6  (1.8-3.6) | 4.0  (2.5-6.3) | 11.3  (7.8-16.2) | 3.6  (2.3-5.5) |
| Female | 3.3  (2.1-5.0) | 2.2  (1.0-4.4) | 0.7  (0.4-1.2) | 1.6  (1.1-2.2) | 0.3  (0.2-0.8) | 2.2  (1.7-3.0) | 2.4  (1.5-3.9) | 7.4  (6.1-8.9) | 2.6  (1.9-3.7) |
| Degree | 2.2  (1.1-4.2) | 0.8  (0.3-2.1) | 0.7  (0.4-1.2) | 1.6  (1.2-2.3) | 0.3  (0.1-0.8) | 0.7  (0.5-1.2) | 2.5  (1.1-5.3) | 11.5  (8.9-14.6) | 2.0  (1.1-3.5) |
| No Degree | 8.1  (5.0-12.8) | 2.6  (1.3-5.0) | 1.0  (0.6-1.7) | 2.4  (1.9-3.1) | 0.7  (0.4-1.4) | 3.5  (2.7-4.4) | 3.4  (2.3-4.9) | 7.4  (5.9-9.3) | 3.3  (2.4-4.4) |
| Age 16-29 | 5.8  (3.7-8.8) |  |  |  |  | 3.6  (2.4-5.5) |  |  | 2.9  (2.2-3.8) |
| Age 30-49 |  | 1.8  (1.0-3.1) |  |  | 0.1  (0.0-0.7) | 1.7  (1.2-2.6) |  |  |  |
| Age 50-66 |  |  | 0.9  (0.6-1.3) | 2.1  (1.8-2.6) | 0.7  (0.4-1.2) | 2.3  (1.7-3.1) | 3.1  (2.2-4.4) | 8.3  (6.9-9.9) |  |

|  | **MCS** | **NS** | **BCS** | **NCDS** | **GS** | **USOC** | **ELSA** | **ALSPAC-G0** | **ALSPAC-G1** |
| --- | --- | --- | --- | --- | --- | --- | --- | --- | --- |
| **Became Employed** | 1.1  (0.7-1.8) | 0.9  (0.4-1.6) | 0.5  (0.3-0.9) | 0.6  (0.4-0.9) | 0.6  (0.3-0.9) | 1.1  (0.8-1.4) | 0.4  (0.2-0.7) | 3.5  (2.7-4.6) | 2.3  (1.6-3.2) |
| Male | 1.3  (0.6-2.8) | 1.0  (0.4-2.3) | 0.2  (0.03-0.7) | 0.8  (0.4-1.3) | 0.2  (0.1-0.8) | 1.1  (0.7-1.8) | 0.2  (0.0-0.6) | 4.0  (2.2-7.1) | 4.5  (2.8-7.0) |
| Female | 1.1  (0.6-2.1) | 0.7  (0.2-1.8) | 0.8  (0.5-1.3) | 0.4  (0.2-0.8) | 0.8  (0.4-1.3) | 1.0  (0.7-1.4) | 0.5  (0.2-1.2) | 3.4  (2.5-4.5) | 1.3  (0.7-2.1) |
| Degree | 1.1  (0.5-1.3) | 0.7  (0.3-1.4) | 0.6  (0.3-1.1) | 0.8  (0.5-1.3) | 0.5  (0.3-1.1) | 1.3  (0.5-1.2) | 0.5  (0.2-1.7) | 4.8  (3.1-7.2) | 3.1  (1.8-5.3) |
| No Degree | 1.0  (0.5-2.4) | 1.0  (0.3-2.3) | 0.5  (0.2-1.0) | 0.5  (0.3-0.9) | 0.6  (0.3-1.3) | 0.9  (0.6-1.4) | 0.3  (0.1-0.7) | 3.2  (2.3-4.4) | 1.9  (1.2-3.0) |
| Age 16-29 | 1.1  (0.7-1.8) |  |  |  | 2.7  (0.5-13.8) | 1.5  (0.9-2.5) |  |  | 2.3  (1.6-3.2) |
| Age 30-49 |  | 0.9  (0.4-1.6) |  |  | 2.5  (0.1-0.9) | 0.9  (0.6-1.5) |  |  |  |
| Age 50-66 |  |  | 0.5  (0.3-0.9) | 0.6  (0.4-0.9) | 0.7  (0.4-1.2) | 0.9  (0.6-1.3) | 0.4  (0.2-0.7) | 3.5  (2.7-4.6) |  |
| **Stable Non-Employed** | 59.6  (54.5-64.4) | 9.5  (7.3-12.3) | 10.4  (9.3-11.5) | 40.0  (38.5-41.4) | 24.5  (22.9-26.2) | 19.2  (18.0-20.4) | 30.0  (27.9-32.2) | 13.7  (11.9-15,8) | 3.3(2.4-4.6) |
| Male | 59.4  (52.5-66.0) | 4.2  (2.2-7.9) | 6.7  (5.4-8.2) | 32.8  (30.8-34.9) | 19.8  (17.3-22.6) | 16.8  (15.0-18.8) | 25.6  (22.5-29.1) | 10.8  (7.6-15.2) | 3.6  (2.3-5.5) |
| Female | 59.9  (55.6-64.0) | 13.7  (10.3-18.0) | 13.8  (12.3-15.4) | 46.9  (44.9-49.0) | 26.8  (24.8-29.0) | 21.3  (19.8-22.9) | 34.0  (31.2-36.9) | 14.6  (12.5-17.0) | 2.6  (1.9-3.7) |
| Degree | 72.0  (66.6-76.9) | 6.5  (4.3-9.7) | 6.5  (5.4-7.8) | 46.8  (44.6-49.0) | 23.4  (21.3-25.8) | 13.7  (12.5-15.0) | 29.1  (25.4-33.1) | 16.0  (12.7-17.0) | 2.0  (1.1-3.5) |
| No Degree | 51.5  (45.7-57.2) | 12.0  (8.6-16.5) | 12.9  (11.4-14.7) | 36.2  (34.3-38.2) | 25.6  (23.3-28.2) | 22.7  (21.0-24.6) | 30.3  (27.8-33.0) | 16.0  (12.7-20.0) | 3.2  (2.4-4.4) |
| Age 16-29 | 59.6  (54.5-64.4) |  |  |  | 21.6  (11.4-37.2) | 22.2  (19.0-25.9) |  |  | 3.3  (2.4-4.6) |
| Age 30-49 |  | 9.5  (7.3-12.3) |  |  | 8.2  (6.5-10.3) | 9.4  (8.0-11.0) |  |  |  |
| Age 50+-66 |  |  | 10.4  (9.3-11.5) | 40.0  (38.5-41.4) | 32.0  (29.8-34.2) | 27.3  (25.6-29.0) | 30.0  (27.9-32.2) | 13.7  (11.9-15.8) |  |

Percentages are weighted (except GS). Note: Analysis for GS, USOC, and ELSA restricted to participants aged 66 and younger.
